# Risk of Bias in Randomized Controlled Trials of Nutrition Interventions for Frailty in Older Adults

**DOI:** 10.1101/2025.08.28.25334686

**Authors:** Malak Mostafa Tawfik, Mark Oremus

## Abstract

We assessed the risk of bias in randomized controlled trials (RCTs) of nutrition-only interventions and holistic frailty outcomes in older adults. We also explored associations between study-level factors and risk of bias. We searched Cochrane, PubMed, and Scopus for published trials between 01/01/2000 and 11/13/2024. Two persons independently screened each citation at the title and abstract, and full text, levels. They also independently conducted data extraction and assessed risk of bias using the Cochrane Risk of Bias 2 tool. We used responses on the tool to develop index scores between 0-1 for each included article, with higher scores indicating lower risk of bias. We regressed the index scores on four study-level factors, i.e., region of publication, year of publication, journal impact factor, and reported use of CONSORT guidelines. Fifteen articles were included in the study: three had low risk of bias, two had some concerns with bias, and ten had high risk of bias. Domain 2 on the Cochrane tool generated the most challenges with bias, largely due to poor reporting of intention-to-treat analysis and lack of information on how this issue might affect trial results. Median index scores were 0.52, 0.53, and 0.86 for articles with high, some concerns, and low risk of bias, respectively (p = 0.0479). However, the index scores were not associated with any study-level factors. Researchers in the field should note potential biases in the design and conduct of RCTs, especially in data analysis and – more specifically – intent-to-treat analysis.

## Introduction

Assessing the risk of bias (RoB) of randomized controlled trials (RCTs) is crucial for evidence-informed decision-making in health policy and practice [1]. While the research community has boosted the integrity of RCTs through mandated pre-trial registration and the CONSORT reporting guidelines [2,3], the design and conduct of clinical trials is left to the diligence and expertise of scientists conducting the work. Thus, published RCTs may appear in a trial registry and adhere to CONSORT, yet still demonstrate high RoB. This situation has led to numerous studies of RoB in RCTs, with the focus being on trials done in specific fields (e.g., inflammatory bowel disease [4], cardiovacular disease [5]), patient groups (e.g., pediatrics [6]), therapies (e.g., tyrosine kinase inhibitors for advanced differentiated thyroid cancer [7]), or geographic regions (e.g., India [8]). By flagging concerns, these studies hope to reduce bias in future trials and improve the evidentiary basis for health decision-making.

### Frailty and nutrition

Frailty is an age-related decline in physiological functioning that increases one’s risk of adverse health outcomes and inhibits a return to normal functioning or homeostasis after ‘minor’ illnesses [9]. Frailty is detrimental to healthy aging and is associated with higher mortality and lower quality-of-life [10,11]. The condition is seen in millions of older adults globally, with prevalence figures ranging from 12-24% [12]. The prevalence and incidence of frailty are expected to increase in conjunction with a projected near doubling of the global older adult population by 2050 [13].

Malnutrition is a major risk factor for frailty because older adults often struggle to consume enough calories to meet their daily energy requirements [14,15]. Many of the risk factors or symptoms of frailty (weight loss or wasting, low muscle mass, low activity and energy levels) are directly related to nutrition [16]. Nutritional interventions can lower the risk of frailty or mitigate frailty-related deterioration (e.g., sarcopenia) [14,17–19].

Nutrition interventions may be a viable alternative to pharmaceutical therapies in the prevention and management of frailty, especially given the association between polypharmacy–a common issue among older adults–and frailty [20–22]. To date, researchers have evaluated many nutrition interventions, including diets, dietary supplements, education, and combinations thereof. Nutrition interventions may sometimes be combined with non-nutrition approaches like exercise [23,24].

### Study objectives

We evaluated the RoB of RCTs investigating at least one ‘clean’ nutrition intervention–not combined with a non-nutrition intervention–for frailty in older adults. Frailty in these trials had to be defined as a holistic construct or syndrome. One such definition is the Fried frailty phenotype, which classifies individuals as frail if they exhibit three or more of the following characteristics: lack of energy (exhaustion), low physical activity levels, weakness (measured using grip strength), shrinking (weight loss), and slow speed (measured by the time to walk 15 feet) [25,26]. Another holistic view of frailty is the deficit accumulation perspective, which sees frailty as a state of ill-health linked to the number of age-related health challenges observed in an older adult [27,28].

Published systematic reviews on nutrition and frailty have found most RCTs suffer from some concerns with bias or high RoB [23,24,29]. However, these reviews mainly included articles that evaluated specific clinical outcomes in persons with frailty (e.g., hand-grip strength, gait speed, fat-free mass), or articles that mixed nutrition and non-nutrition interventions in the same treatment groups. Since frailty is a condition featuring a constellation of health challenges, we explored RoB in RCTs that investigated the efficacy of nutrition interventions on holistic frailty outcomes. We developed a process to quantify our RoB assessments on the Cochrane Risk of Bias 2 tool (CRoB2) [30] and explore whether study-level factors such as year of publication were associated with RoB. We did not conduct a systematic review and undertook this study to help promote the design and conduct of optimal trials in nutrition and frailty.

## Materials and Methods

### Reporting

Reporting was done in accordance with the Preferred Reporting Items for Systematic Review and Meta-Analysis extension for scoping reviews (PRISMA-ScR) [31] (Supplementary appendix 1), based on the recommendation of the ‘Right Review’ tool [32].

### Literature Search

We searched Cochrane Central Register of Controlled Trials, PubMed, and Scopus for RCTs published in peer reviewed journals from 2000 (January 01) to 2024 (November 13). The search strategy (Supplementary appendix 2) was developed with the aid of a medical librarian. We also checked the reference lists of included articles to identify additional citations for consideration.

### Eligibility Criteria

We screened citations for inclusion based on the PICOTS criteria. <u>Population</u>: Articles had to include results for participants aged ≥ 60 years (older adults) who received a frailty classification (non frail, pre-frail, frail, etc.) using any clinically accepted measure of frailty [11], provided frailty was assessed holistically. This allowed us to include prevention-focused RCTs in addition to trials of frailty management. <u>Intervention and comparator</u>: At least one trial arm had to contain any type of solo nutrition intervention (e.g., food diet, dietary supplements, education about healthy eating choices). Any comparator was acceptable, including the same nutrition intervention if it was part of a multimodal approach (e.g., a combined diet and physical activity program versus diet alone). <u>Outcome</u>: We considered any holistic measure of frailty, such as the Fried frailty phenotype or deficit accumulation perspective, and assessed RoB on this outcome. <u>Time</u>: We accepted any length of follow-up. <u>Setting</u>: RCTs of older adults in any setting, e.g., community dwelling, hospitalized, institutionalized, etc. were eligible for inclusion.

### Screening

We conducted title and abstract, and full text, screening using Covidence systematic review software (Veritas Health Innovation, Melbourne, Australia). Two screeners independently evaluated the titles and abstracts of the retrieved citations against the eligibility criteria and promoted articles to the full text phase if they met or maybe met all criteria. At the full text level, the two screeners independently read the full articles and promoted reports that met all criteria to data extraction. Any disagreements throughout the screening process were resolved by consensus or by a third team member who was not one of the initial screeners.

### Data Extraction

Two screeners independently conducted data extraction and disagreements were resolved as described above. Two data extraction forms were produced, one to collect article information and the other based on the CRoB2 tool. Article information included first author, year of publication, region of publication (Asia or other), journal impact factor at the time of publication [33], reported use of CONSORT guidelines (yes or no), and whether the authors reported sources of funding (yes or no). We categorized region as ‘Asia’ or ‘other’ because most included articles reported on research conducted in Asia; the number of non-Asian articles was too small and too widely distributed geographically to support more than two response levels in regression analysis. For reporting of funding sources, we gave ‘no’ responses when authors simply declared an absence of funding conflicts or did not provide any information in this area; articles received ‘yes’ responses when authors reported the names of funding agencies, organizations, etc., or clearly specified that no funding was received to conduct the trial. RoB assessments followed existing guidance for the CRoB2 tool [34].

### Data Analysis

To investigate whether year of publication, region of publication, journal impact factor, reported use of CONSORT guidelines, and reported sources of funding were associated with RoB, we piloted the use of an index score ranging from 0-1, with higher scores representing lower levels of bias. To compute the index score, we assigned a numerical value to each CRoB2 question response, as follows: (i) if Y (yes) answers indicated low risk of bias, then they were assigned scores of Y = 1 and PY (probably yes) = 0.5; (ii) if Y (yes) answers indicated high risk of bias, then they were assigned scores of Y = -1 and PY (probably yes) = -0.5; (iii) we applied the same logic to N (no) and PN (probably no) responses; (iv) NI (no information) answers were scored 0; and (v) NA (not applicable) answers were left blank. Supplementary appendix 3 shows whether Y or N answers indicated low RoB for each CRoB2 domain. An article’s index score was calculated by summing together the numerical values for the CRoB2 responses and dividing them by the total number of responses that received anything other than an NA answer. We then regressed the index scores on year of publication, region of publication, journal impact factor, reported use of CONSORT guidelines, and reported sources of funding.

We used R v4.3.3 (The R Foundation for Statistical Computing, Vienna, Austria) and the ‘lm’ function to conduct a multivariable linear regression exploring whether study-level factors were associated with index scores. We employed R’s ‘robvis’ package [35] to produce plots showing the (i) distribution of RoB assessments in each CRoB2 domain and (ii) RoB scores for each included article. We also utilized R’s ‘gglot2’ and ‘ggpubr’ packages [36,37] to generate graphs. The level of statistical significance was α = 0.05. No ethics approval was required for this research because we relied solely on the contents of published articles.

## Results

### Article selection process

The literature search identified 1,253 citations, of which 809 moved to title and abstract screening after removing duplicates. One hundred seventy articles were assessed at full text screening and 15 were included in our study. Figure 1 summarizes the flow of studies through the screening process.

**Figure 1.**
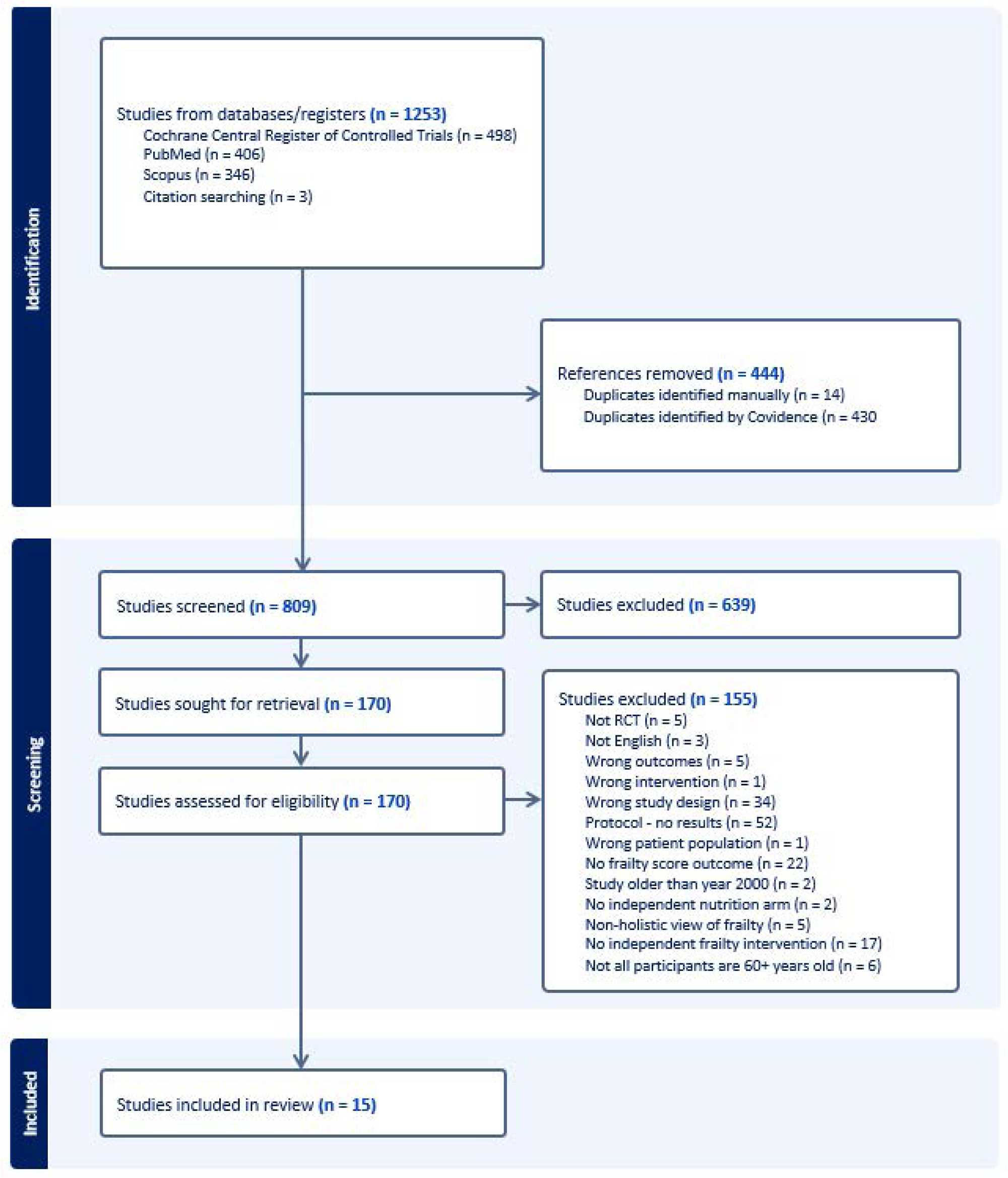
Flow diagram of the study selection process.

### Characteristics of included articles

The characteristics of the 15 included articles are shown in Table 1 [38–52]. They were published between 2013 and 2023 and impact factors ranged from 1.4 to 13.1. The impact factor for one article was unavailable [47]. This same article did not report funding sources, whereas the other articles did report these sources. Only 4 of 15 articles reported using the CONSORT guidelines [39,44,46,50].

**Table 1.** Article characteristics.

| **Study** | **Region of Publication** | **Journal Impact Factor** | **Reported Use of CONSORT Guidelines** | **Reported Funding Sources** | **Risk of Bias** | **Index Score** |
| --- | --- | --- | --- | --- | --- | --- |
| Badrasawi, 2016 [38] | Other | 2.5810 | No | Yes | High | 0.8571 |
| Biesek, 2021 [39] | Asia | 4.6140 | Yes | Yes | High | 0.4722 |
| Buigues, 2016 [40] | Asia | 3.2260 | No | Yes | High | 0.6471 |
| Cai, 2022 [41] | Asia | 4.1000 | No | Yes | Low | 1.0000 |
| Chatterjee, 2018 [42] | Other | 2.8180 | No | Yes | High | 0.1250 |
| Gagesch, 2023 [43] | Asia | 3.9000 | No | Yes | High | 0.6667 |
| Hsieh, 2019 [44] | Other | 6.7140 | Yes | Yes | High | 0.2500 |
| Hutchins-Wiese, 2013 [45] | Asia | 2.6590 | No | Yes | High | 0.6429 |
| Kim, 2015 [46] | Other | 3.0570 | Yes | Yes | Low | 0.8571 |
| Na, 2021 [47] | Other | NA | No | No | High | 0.5625 |
| Ng, 2015 [48] | Other | 5.6100 | No | Yes | Some | 0.5313 |
| Park, 2018 [49] | Other | 6.5680 | No | Yes | Low | 0.8571 |
| Park, 2023 [50] | Other | 3.5000 | Yes | Yes | Some | 0.5313 |
| Teh, 2022 [51] | Asia | 13.1000 | No | Yes | High | 0.4118 |
| Wu, 2018 [52] | Other | 1.3750 | No | Yes | High | 0.2632 |

Abbreviations: CONSORT, Consolidated Standards of Reporting Trials; NA, not available.

### Risk of bias

Of the 15 articles, 3 had low risk of bias [41,46,49], 2 had some concerns with bias [48,50], and 10 had high risk of bias [38–40,42–45,47,51,52]. Figure 2 visualizes RoB in each domain and overall; Figure 3 visualizes RoB in each article. Domain 3 (RoB “due to missing outcome data”) and Domain 5 (ROB due to “selection of the reported results”) had the lowest overall risk of bias across all included articles. Domain 2 (RoB “due to deviations from intended intervention”) showed the highest risk of bias, with 5 articles [42,44,47,48,50] rated ‘some concerns’ and 7 articles [38–40,43,45,51,52] rated ‘high’ RoB. The three articles rated ‘low’ RoB under Domain 2 were also the only articles to receive ‘low’ overall RoB rankings [41,46,49].

**Figure 2.**
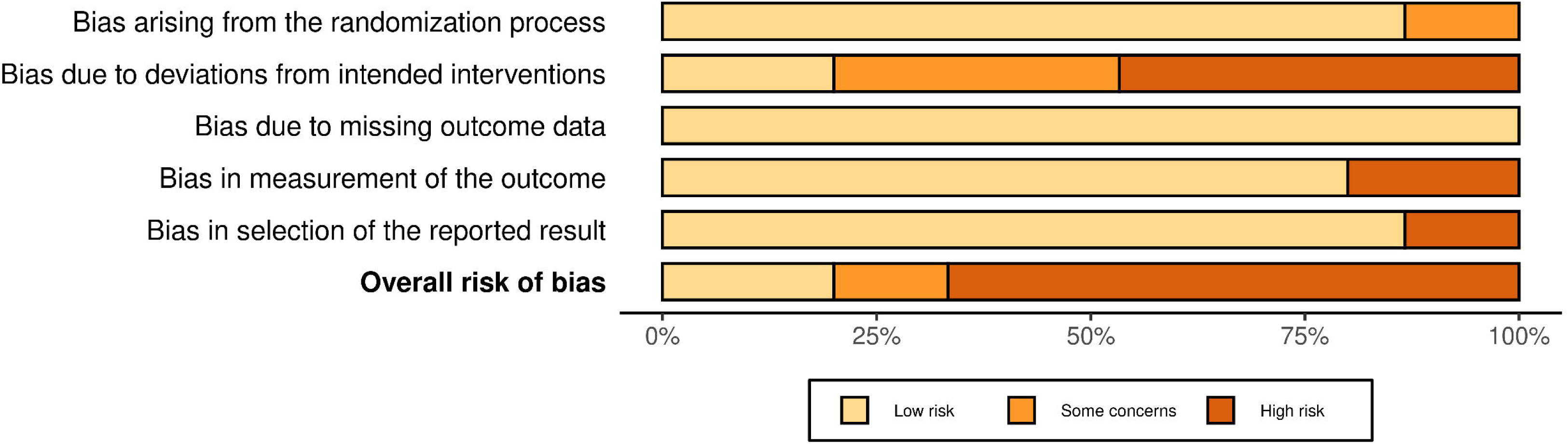
CRoB2 summary graph for the included studies (n = 16) by domain.

**Figure 3.**
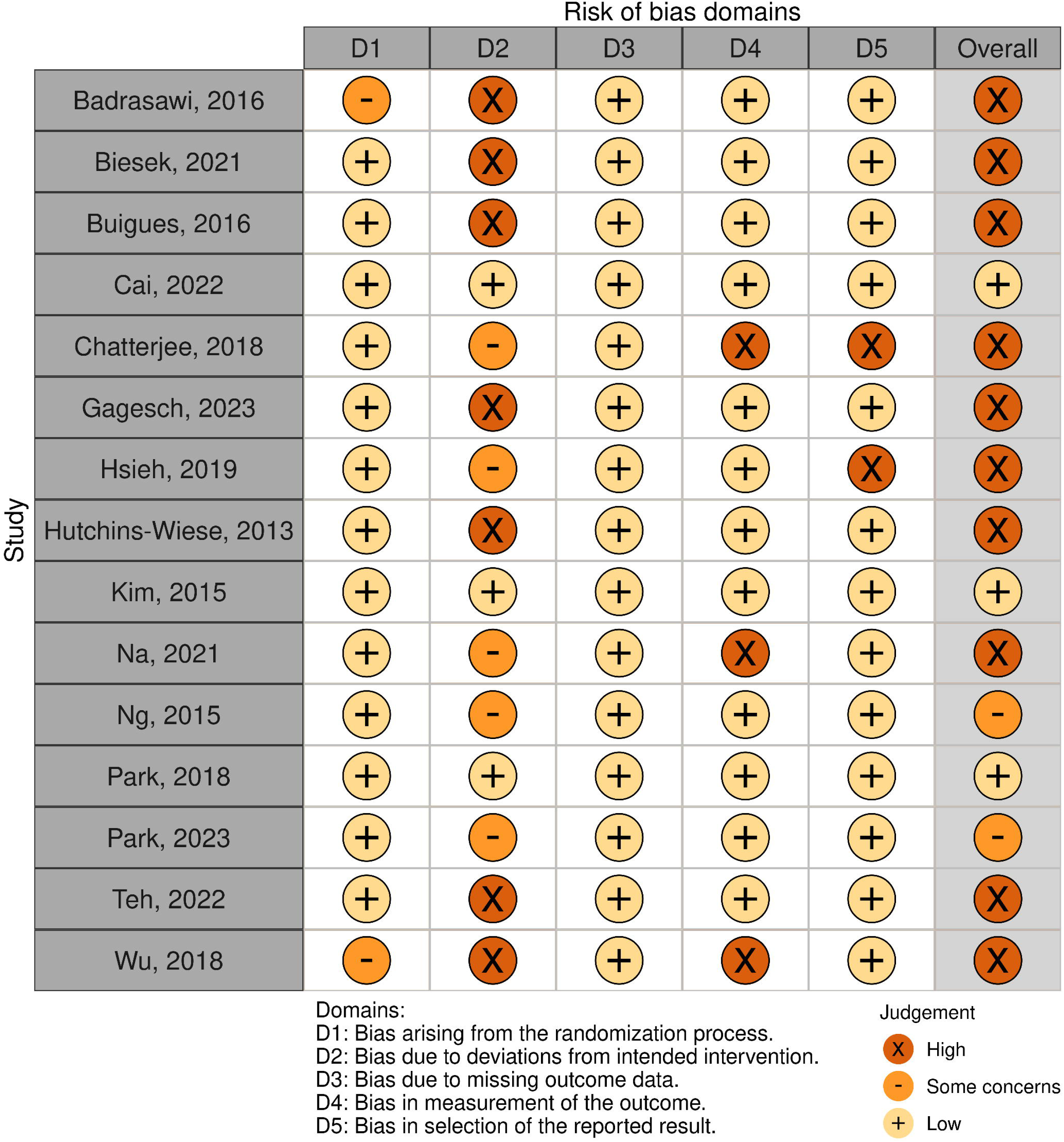
CRoB2 summary graph for the included studies (n = 16) by article.

One [42] of the two trials that showed high RoB in Domain 5 was retrospectively entered in a clinical trial registry and the other was not entered in any registry [45]. One further trial was entered retrospectively [44]. Four trials [38,40,45,47] were not entered into any clinical trial registry and none of these four reported use of CONSORT.

### Index scores and multivariable regression

The translation of responses on the CRoB2 tool into index scores is shown in Supplementary appendix 3. Figure 4 shows the index scores were slightly left skewed. The median index score was 0.56 (interquartile range: 0.46) and the range was 0.13 to 1.00 (Table 1). The median index scores for high, some concerns, and low RoB were 0.52, 0.53, and 0.86, respectively (Kruskal-Wallis: *X*^2^ [2, n = 15] = 6.08, p = 0.0479; Supplementary appendix 4).

**Figure 4.**
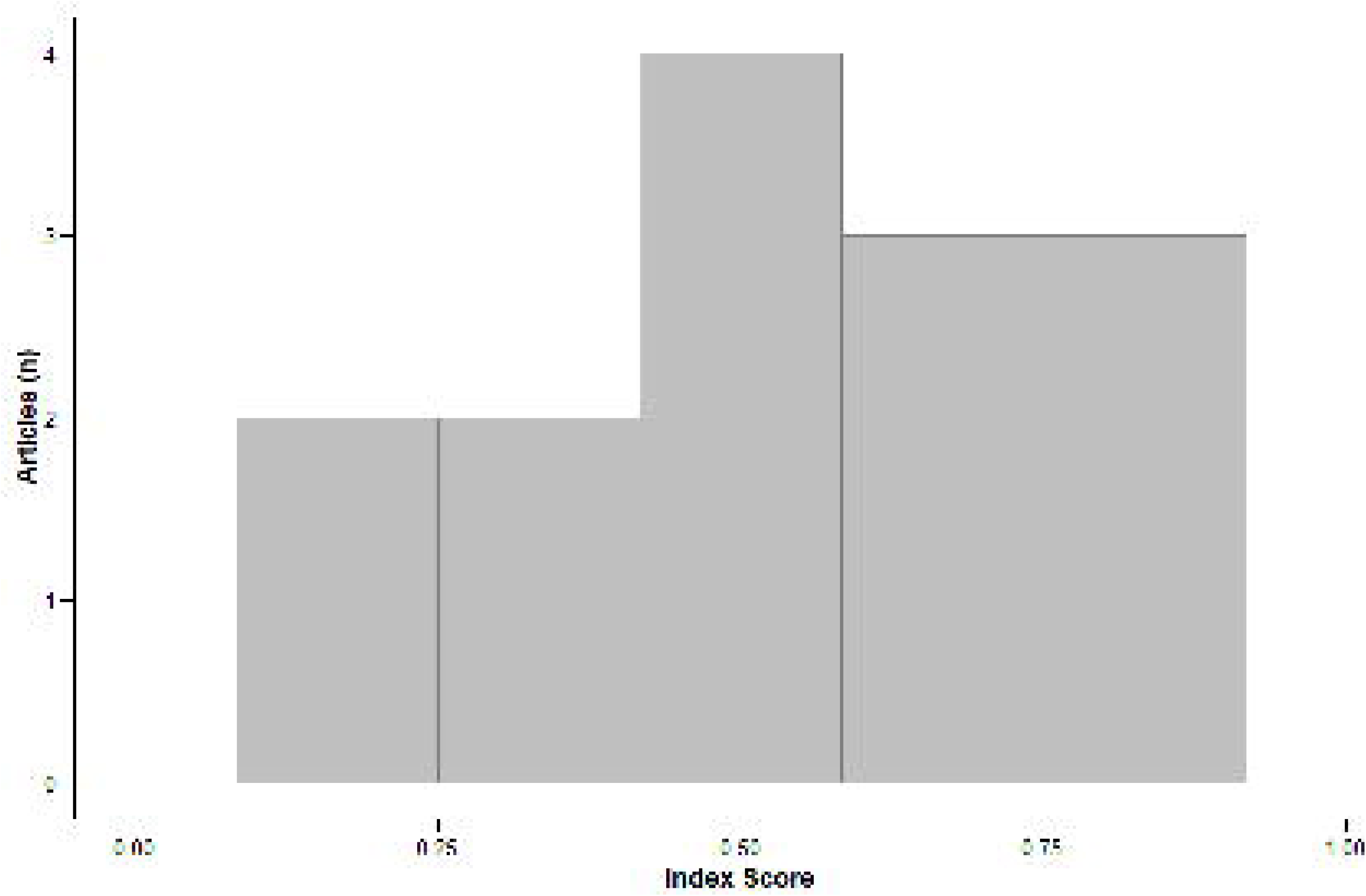
Distribution of index scores.

For the multivariable regression analysis, we excluded the article for which journal impact factor was unavailable [47]. Since the remaining 14 articles reported funding sources (‘yes’ response), we excluded this variable from the model. The remaining four study-level variables were not associated with the index scores (Table 2).

**Table 2.** Multiple linear regression.

| **Variable** | **Regression Coefficient (95% CI)** |
| --- | --- |
| Region of Publication | 0.132 (-0.269, 0.532) |
| Year of Publication | -0.010 (-0.074, 0.056) |
| Journal Impact Factor | -0.013 (-0.083, 0.057) |
| Reported Use of CONSORT Guidelines | -0.029 (-0.457, 0.400) |

Abbreviations: CI, confidence interval; CONSORT, Consolidated Standards of Reporting Trials.

## Discussion

### Major risk of bias challenges

Of the 15 included articles, all except 3 showed some concerns or high risk of bias. The major RoB challenge was Domain 2, where 12 of 15 articles scored some concerns or high RoB. For 5 of these 12 articles [39,40,43,45,51], the RoB level on the other four domains was low. Other reviews of nutrition interventions for frailty that rated RoB using CRoB2 found similar results. de Moraes et al. rated only 5 of 19 RCTs [24], and Sirikul et al. rated 13 of 24 RCTs [29], with low overall RoB. Domain 2 was the poorest performing bias domain in de Moraes et al. and the second poorest in Sirikul et al. (Figure 5).

**Figure 5.**
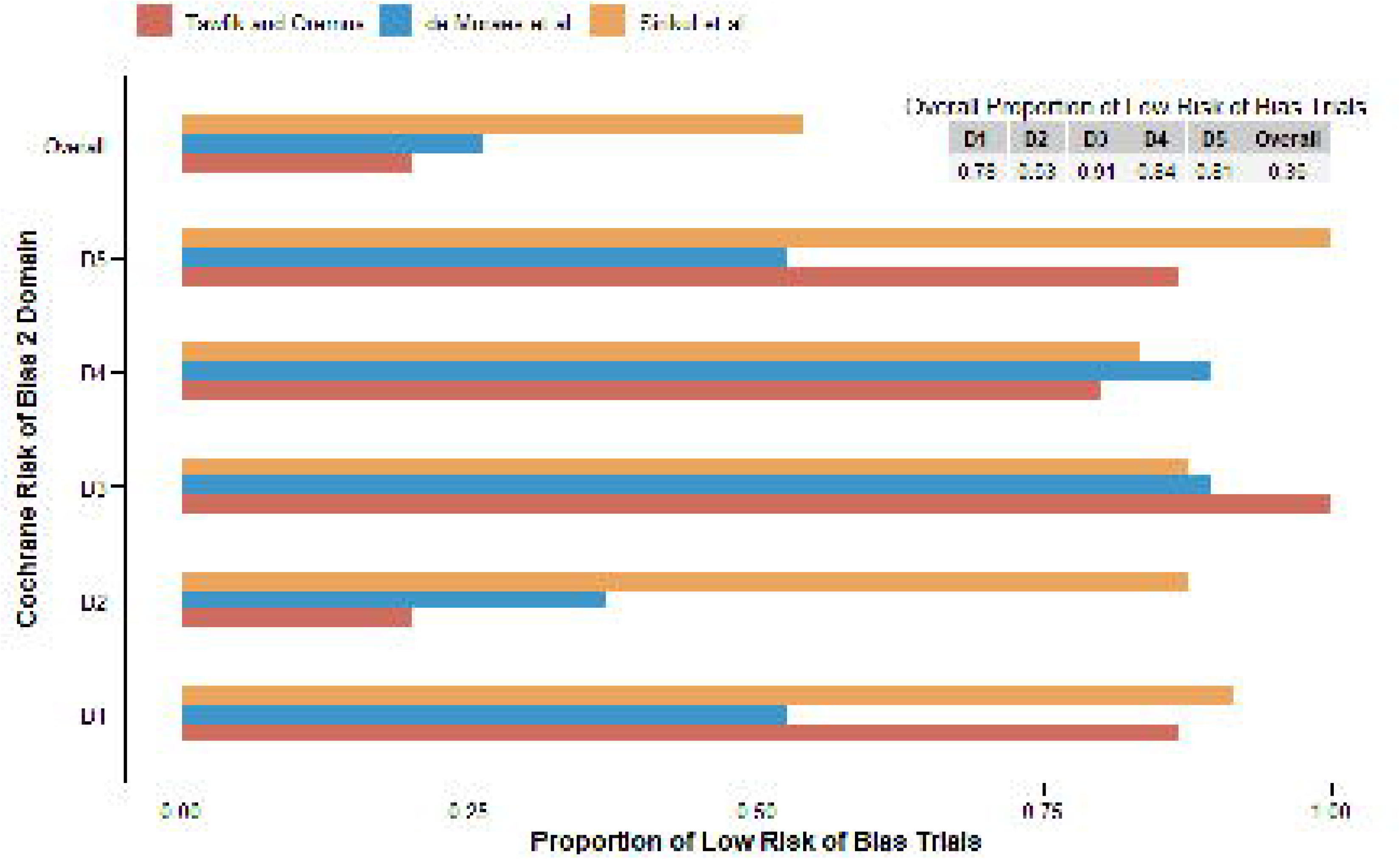
Randomized controlled trials with low risk of bias in nutrition and frailty.

We assessed the effect of assignment to intervention in Domain 2 and found the major concerns rested with questions 2.6 and 2.7 on CRoB 2. Question 2.6 asks whether an appropriate analysis such as intention-to-treat (ITT) or modified ITT was used to estimate the effect of assignment to intervention. Eight articles contained no information on the topic [38– 40,43,45,47,50,52] and one was rated ‘no’ [51] (Supplementary appendix 3). Of these 9 articles, we had insufficient information in the case of 6 [38–40,43,45,52] to assess whether inappropriate analyses had an adverse impact on the results (question 2.7). These question responses, combined with the remaining components of the algorithm to rate the level of bias in Domain 2 [34], led to poor performance on this domain.

The importance of ITT analysis of RCT data was raised decades ago [53,54] and is a well-known pillar of trial methodology [55,56]. Given the publication dates of the articles, we expected the authors would be aware of the need for ITT analysis and suspected the issue could revolve around poor reporting of methods, rather than poor conduct of analyses. Of the 12 articles scoring some concerns or high RoB on this domain, 9 [38,40,42,43,45,47,48,51,52] did not report the use of CONSORT. Differences between the reporting of RCTs and what was actually done in the conduct of trials is a major criticism of assessing RoB using published trial reports [57]. The authors of some of the included articles may have followed proper analytical procedures and improperly reported what they did.

To address RoB from inappropriate analysis methods, trial researchers in nutrition and frailty should utilize and explicitly describe their ITT or modified ITT approaches. Item 21 of the CONSORT guidelines calls for such explicitness [3]. Researchers should also avoid analysis methods that exclude or reassign participants following randomization, save for participants with missing data [55,56].

### Study-level factors and risk of bias index scores

None of the study-level factors shown in Table 2 were statistically significantly associated with the index scores. This was likely due to a lack of power (n = 14 articles). Explorations of study-level factors associated with RoB can be frustrated when a small number of trials are included in systematic reviews or meta-analyses. Published reviews include a median of 15 articles [58], which is insufficiently large to achieve statistical power in most regression analyses. While reviews of the methodological quality and RoB of 100,000+ trials, published over decades [59], are helpful to assess research trends and improve the scientific enterprise, most researchers will want to focus on promoting bias minimization in their specific fields.

We created the index scores to explore the impact of study-level factors on RoB in RCTs of nutrition interventions for frailty. Quantifying RoB generally does not occur because of criticisms against the practice [60–62]. However, we wanted to test a process that resembled meta-regression, which seeks to identify study-level factors associated with effect sizes in meta-analyses. Such a process would be an alternative to the bivariate comparisons [63] and counts [64] typically employed to examine associations between study-level factors and RoB. These latter analytical approaches do not account for the independent effect of each study-level factor after controlling for all other factors.

On the surface, the index scores appeared to discriminate between high, some concerns, and low RoB trials. In accordance with Viswanathan et al. [65], we provided specific rules for computing the index scores and recognize these scores are “subjective and arbitrary [p. 32]” [65]. We also did not assign relative values to the scores, e.g., a trial with an index score of 0.70 was not viewed as being in the upper range of RCTs with some concerns about bias, compared to studies with index scores from 0.50 to 0.60.

We could have treated RoB as a categorical outcome in the regression analysis. With only two articles in the ‘some concerns’ category, combination with the ‘high’ RoB group would have been likely. The validity of combining two arguably different levels of bias is questionable. Had a larger number of articles been available, ordinal or multinomial regression may have been possible. These analyses come with caveats that could affect implementation. For instance, alternative strategies are required when one or more independent variables do not meet the proportional odds assumption for ordinal regression, and complexities arise when reporting and summarizing multinomial regression results involving many categorical variables.

### Strengths and limitations

To maximize our ability to obtain all relevant RCTs on nutritional interventions for frailty, we followed PRISMA-ScR guidelines and conducted a detailed literature search across three databases. To promote the validity of the study, two independent screeners assessed the articles at two levels (title and abstract, full text), conducted RoB assessments using the CRoB2 tool, and extracted study information. Although an earlier meta-epidemiological study examined RoB in 927 nutrition RCTs [66], this work did not include trials with holistic frailty outcomes. Large meta-epidemiological studies may not apply to key research challenges in sub-domains of a field of inquiry [67].

Our study is limited by the small sample size, which led to low power to assess study-level factors that might be associated with RoB. The results of our study only apply to RCTs that evaluated at least one independent nutrition intervention against a holistic outcome. The study was not a full systematic review because the intent was to explore RoB in a specific set of nutrition trials.

## Conclusion

As more research is conducted to investigate nutritional interventions for frailty, researchers must consider potential bias threats in the design and conduct of RCTs. We hope additional tools will become available in this regard, such as improved methods to assess study-level factors associated with RoB.

## Data Availability

All data are freely available from the corresponding author. The R code and datasets for median index scores, Kruskal-Wallis test, figures 4 and 5, regression in table 2, and Supplementary appendix 4 are available on GitHub (https://github.com/moremus/Tawfik_Oremus.git).

## Acknowledgements

The authors wish to thank Paniz Haghighi, Haya Abu Ghanem, and Asher C. Wiggans for serving as screeners.

## CRediT Authorship Contribution Statement

**Malak Mostafa Tawfik**: Conceptualization, Data curation, Formal analysis, Investigation, Software, Visualization, Writing – original draft; **Mark Oremus:** Conceptualization, Formal analysis, Methodology, Project administration, Supervision, Validation, Visualization, Writing – review and editing. All authors read and approved the version of the manuscript being submitted

## Ethical Approval

Ethics approval was not required for this study because all the information came from published articles. No data from individual persons were used in this study.

## References

1. Phillips MR, Kaiser P, Thabane L, Bhandari M, Chaudhary V. Risk of bias: why measure it, and how? Eye 2022;36:346–348. 10.1038/s41433-021-01759-9.

2. Elliott TR. Registering randomized clinical trials and the case for CONSORT. Exp Clin Psychopharmacol 2007;15:511–518. 10.1037/1064-1297.15.6.511.

3. Hopewell S, Chan A-W, Collins GS, Hróbjartsson A, Moher D, Schulz KF, et al. CONSORT 2025 explanation and elaboration: updated guideline for reporting randomised trials. BMJ 2025:e081124. 10.1136/bmj-2024-081124.

4. Gordon M, Khudr J, Sinopoulou V, Lakunina S, Rane A, Akobeng A. Quality of reporting inflammatory bowel disease randomised controlled trials: a systematic review. BMJ Open Gastroenterol 2024;11:e001337. 10.1136/bmjgast-2023-001337.

5. Baasan O, Freihat O, Nagy DU, Lohner S. Change over five years in important measures of methodological quality and reporting in randomized cardiovascular clinical trials. J Cardiovasc Dev Dis 2023;11:2. 10.3390/jcdd11010002.

6. Huo B, Xu S, Liu Y, Su L, Jia Y, Ai M, et al. Quality assessment of paediatric randomized controlled trials published in China from 1999 to 2022: a cross-sectional study. BMC Pediatr 2024;24:364. 10.1186/s12887-024-04839-3.

7. Dedivitis RA, Castro MAF de, Boni AMD, Alvares ACB, Tresso AJP, Oliveira AD de, et al. The methodological and reporting quality of randomized controlled trials of tyrosine kinase inhibitors for advanced differentiated thyroid cancer: meta-research study. Head Neck 2024;46:1683–1697. 10.1002/hed.27679.

8. Patil SR, Bhide SS. Assessment of risk of bias in randomized controlled trials published in Indian journals pertaining to pharmacology. Perspect Clin Res 2023;14:16–19. 10.4103/picr.PICR_19_21.

9. Won CW. Up-to-date knowledge of frailty. J Korean Med Assoc 2022;65:108–114.

10. Statistics Canada. Association of frailty and pre-frailty with increased risk of mortality among older Canadians. 2021. https://www150.statcan.gc.ca/n1/pub/82-003-x/2021004/article/00002-eng.htm.

11. Kim DH, Rockwood K. Frailty in older adults. N Engl J Med 2024;391:538–548. 10.1056/NEJMra2301292.

12. Dlima SD, Hall A, Aminu AQ, Akpan A, Todd C, Vardy ERLC. Frailty: a global health challenge in need of local action. BMJ Glob Health 2024;9. 10.1136/bmjgh-2024-015173.

13. World Health Organization. Ageing and health. 2024. https://www.who.int/news-room/fact-sheets/detail/ageing-and-health.

14. Buhl SF, Beck AM, Olsen PØ, Kock G, Christensen B, Wegner M. Relationship between physical frailty, nutritional risk factors and protein intake in community-dwelling older adults. Clin Nutr ESPEN 2022;49:449–458.

15. Lorenzo-López L, Maseda A, Labra C, Regueiro-Folgueira L, Rodríguez-Villamil JL, Millán-Calentí JC. Nutritional determinants of frailty in older adults: a systematic review. BMC Geriatr 2017;17:108. 10.1186/s12877-017-0496-2.

16. Lochlainn MN, Cox NJ, Wilson T, Hayhoe RPG, Ramsay SE, Granic A. Nutrition and frailty: opportunities for prevention and treatment. Nutrients 2021;13:2349.

17. Morley JE. Nutrition and aging well. J Am Med Dir Assoc 2017;18:91–94.

18. Rahi B, Colombet Z, Harmand MGC, Dartigues JF, Boirie Y, Letenneur L. Higher protein but not energy intake is associated with a lower prevalence of frailty among community-dwelling older adults in the French Three-City cohort. J Am Med Dir Assoc 2016;17:672.e7–672.e11. 10.1016/j.jamda.2016.05.005.

19. Jeejeebhoy KN. Malnutrition, fatigue, frailty, vulnerability, sarcopenia and cachexia. Curr Opin Clin Nutr Metab Care 2012;15:213–219.

20. Daimaru K, Osuka Y, Kojima N, Mizukami K, Motokawa K, Iwasaki M, et al. Associations of polypharmacy with frailty severity and each frailty phenotype in community-dwelling older adults: Itabashi Longitudinal Study on Aging. Geriatr Gerontol Int 2024;24:196–201. 10.1111/ggi.14789.

21. Gutiérrez-Valencia M, Izquierdo M, Cesari M, Casas-Herrero Á, Inzitari M, Martínez-Velilla N. The relationship between frailty and polypharmacy in older people: A systematic review. Br J Clin Pharmacol 2018;84:1432–1444. 10.1111/bcp.13590.

22. Saum K-U, Schöttker B, Meid AD, Holleczek B, Haefeli WE, Hauer K, et al. Is polypharmacy associated with frailty in older people? Results from the ESTHER cohort study. J Am Geriatr Soc 2017;65:e27–e32. 10.1111/jgs.14718.

23. Li W, Wu Z, Liao X, Geng D, Yang J, Dai M, et al. Nutritional management interventions and multi-dimensional outcomes in frail and pre-frail older adults: a systematic review and meta-analysis. Arch Gerontol Geriatr 2024;125:105480. 10.1016/j.archger.2024.105480.

24. de Moraes MB, Avgerinou C, Fukushima FB, Vidal EIO. Nutritional interventions for the management of frailty in older adults: systematic review and meta-analysis of randomized clinical trials. Nutr Rev 2021;79:889–913. 10.1093/nutrit/nuaa101.

25. Fried LP, Cohen AA, Xue Q-L, Walston J, Bandeen-Roche K, Varadhan R. The physical frailty syndrome as a transition from homeostatic symphony to cacophony. Nat Aging 2021;1:36–46. 10.1038/s43587-020-00017-z.

26. Fried LP, Tangen CM, Walston J, Newman AB, Hirsch C, Gottdiener J. Frailty in older adults: evidence for a phenotype. J Gerontol Biol Sci Med Sci 2001;56:M146–56.

27. Howlett SE, Rutenberg AD, Rockwood K. The degree of frailty as a translational measure of health in aging. Nat Aging 2021;1:651–665. 10.1038/s43587-021-00099-3.

28. Mitnitski AB, Mogilner AJ, Rockwood K. Accumulation of deficits as a proxy measure of aging. ScientificWorldJournal 2001;1:323–336. 10.1100/tsw.2001.58.

29. Sirikul W, Buawangpong N, Pinyopornpanish K, Siviroj P. Impact of multicomponent exercise and nutritional supplement interventions for improving physical frailty in community-dwelling older adults: a systematic review and meta-analysis. BMC Geriatr 2024;24:958. 10.1186/s12877-024-05551-8.

30. Sterne JAC, Savović J, Page MJ, Elbers RG, Blencowe NS, Boutron I, et al. RoB 2: a revised tool for assessing risk of bias in randomised trials. BMJ 2019;366:l4898. 10.1136/bmj.l4898.

31. Tricco AC, Lillie E, Zarin W, O’Brien KK, Colquhoun H, Levac D, et al. PRISMA Extension for Scoping Reviews (PRISMA-ScR): checklist and explanation. Ann Intern Med 2018;169:467–473. 10.7326/M18-0850.

32. Amog K, Pham B, Courvoisier M, Mak M, Booth A, Godfrey C, et al. The web-based “Right Review” tool asks reviewers simple questions to suggest methods from 41 knowledge synthesis methods. J Clin Epidemiol 2022;147:42–51. 10.1016/j.jclinepi.2022.03.004.

33. Clarivate. Journal impact factors. J Cit Rep. 2025. https://jcr-clarivate-com.proxy.lib.uwaterloo.ca/jcr/home.

34. Higgins JPT, Savović J, Page MJ, Elbers RG, Sterne J. Chapter 8: Assessing risk of bias in a randomized trial [last updated October 2019]. In: Higgins JPT, Thomas J, Chandler J, Cumpston M, Li T, Page MJ, Welch VA (eds). Cochrane Handb. Syst. Rev. Interv. Version 65, Cochrane; 2024.

35. McGuinness L. robvis: An R package and web application for visualising risk-of-bias Assessments. 2025. https://github.com/mcguinlu/robvis.

36. Wickham H. Ggplot2: Elegant Graphics for Data Analysis. 2nd ed. Springer International Publishing, Cham, Switzerland, 2016.

37. Kassambara A. ggpubr: “ggplot2” based publication ready plots. R package version 0.6.0.999. 2025. https://github.com/kassambara/ggpubr.

38. Badrasawi M, Shahar S, Zahara AM, Nor Fadilah R, Singh DKA. Efficacy of L-carnitine supplementation on frailty status and its biomarkers, nutritional status, and physical and cognitive function among prefrail older adults: a double-blind, randomized, placebo-controlled clinical trial. Clin Interv Aging 2016;11:1675–1686. 10.2147/CIA.S113287.

39. Biesek S, Vojciechowski AS, Filho JM, Menezes Ferreira ACR de, Borba VZC, Rabito EI, et al. Effects of exergames and protein supplementation on body composition and musculoskeletal function of prefrail community-dwelling older women: a randomized, controlled clinical trial. Int J Environ Res Public Health 2021;18. 10.3390/ijerph18179324.

40. Buigues C, Fernández-Garrido J, Pruimboom L, Hoogland AJ, Navarro-Martínez R, Martínez-Martínez M, et al. Effect of a prebiotic formulation on frailty syndrome: a randomized, double-blind clinical trial. Int J Mol Sci 2016;17. 10.3390/ijms17060932.

41. Cai Y, Wanigatunga AA, Mitchell CM, Urbanek JK, Miller ER 3rd, Juraschek SP, et al. The effects of vitamin D supplementation on frailty in older adults at risk for falls. BMC Geriatr 2022;22:312. 10.1186/s12877-022-02888-w.

42. Chatterjee P, Kumar P, Kandel R, Madan R, Tyagi M, Kumar DA, et al. Nordic walking training and nutritional supplementation in pre-frail older Indians: an open-labelled experimental pre-test and post-test pilot study to develop intervention model. BMC Geriatr 2018;18:212. 10.1186/s12877-018-0890-4.

43. Gagesch M, Wieczorek M, Vellas B, Kressig RW, Rizzoli R, Kanis J, et al. Effects of vitamin D, omega-3 fatty acids and a home exercise program on prevention of pre-frailty in older adults: the DO-HEALTH randomized clinical trial. J Frailty Aging 2023;12:71–77. 10.14283/jfa.2022.48.

44. Hsieh T-J, Su S-C, Chen C-W, Kang Y-W, Hu M-H, Hsu L-L, et al. Individualized home-based exercise and nutrition interventions improve frailty in older adults: a randomized controlled trial. Int J Behav Nutr Phys Act 2019;16:119. 10.1186/s12966-019-0855-9.

45. Hutchins-Wiese HL, Kleppinger A, Annis K, Liva E, Lammi-Keefe CJ, Durham HA, et al. The impact of supplemental n-3 long chain polyunsaturated fatty acids and dietary antioxidants on physical performance in postmenopausal women. J Nutr Health Aging 2013;17:76–80. 10.1007/s12603-012-0415-3.

46. Kim H, Suzuki T, Kim M, Kojima N, Ota N, Shimotoyodome A, et al. Effects of exercise and milk fat globule membrane (MFGM) supplementation on body composition, physical function, and hematological parameters in community-dwelling frail Japanese women: a randomized double blind, placebo-controlled, follow-up trial. PloS One 2015;10:e0116256. 10.1371/journal.pone.0116256.

47. Na W, Kim J, Kim H, Lee Y, Jeong B, Lee SP, et al. Evaluation of oral nutritional supplementation in the management of frailty among the elderly at facilities of community care for the elderly. Clin Nutr Res 2021;10:24–35. 10.7762/cnr.2021.10.1.24.

48. Ng TP, Feng L, Nyunt MSZ, Feng L, Niti M, Tan BY. Nutritional, physical, cognitive, and combination interventions and frailty reversal among older adults: a randomized controlled trial. Am J Med 2015;128:1225.e1–1236.e1.

49. Park Y, Choi J-E, Hwang H-S. Protein supplementation improves muscle mass and physical performance in undernourished prefrail and frail elderly subjects: a randomized, double-blind, placebo-controlled trial. Am J Clin Nutr 2018;108:1026–1033. 10.1093/ajcn/nqy214.

50. Park W, Lee J, Hong K, Park H-Y, Park S, Kim N, et al. Protein-added healthy lunch-boxes combined with exercise for improving physical fitness and vascular function in pre-frail older women: a community-based randomized controlled trial. Clin Interv Aging 2023;18:13–27. 10.2147/CIA.S391700.

51. Teh R, Barnett D, Edlin R, Kerse N, Waters DL, Hale L, et al. Effectiveness of a complex intervention of group-based nutrition and physical activity to prevent frailty in pre-frail older adults (SUPER): a randomised controlled trial. Lancet Healthy Longev 2022;3:e519–e530. 10.1016/S2666-7568(22)00124-6.

52. Wu SY, Hsu LL, Hsu CC, Hsieh TJ, Su SC, Peng YW, et al. Dietary education with customised dishware and food supplements can reduce frailty and improve mental well-being in elderly people: a single-blind randomized controlled study. Asia Pac J Clin Nutr 2018;27:1018–1030. 10.6133/apjcn.032018.02.

53. Armitage P. The construction of comparable groups. In: Hill AB (ed). Controlled Clinical Trials. Blackwell, Oxford, 1960. pp. 14–18.

54. Montori VM, Guyatt GH. Intention-to-treat principle. CMAJ Can Med Assoc J 2001;165:1339–1341.

55. Chalmers I, Matthews R, Glasziou P, Boutron I, Armitage P. Trial analysis by treatment allocated or by treatment received? Origins of ‘the intention-to-treat principle’ to reduce allocation bias: Part 1. J R Soc Med 2023;116:343–350. 10.1177/01410768231203922.

56. Chalmers I, Matthews R, Glasziou P, Boutron I, Armitage P. Trial analysis by treatment allocated or by treatment received? Origins of ‘the intention-to-treat principle’ to reduce allocation bias: Part 2. J R Soc Med 2023;116:386–394. 10.1177/01410768231203936.

57. Soares HP, Daniels S, Kumar A, Clarke M, Scott C, Swann S, et al. Bad reporting does not mean bad methods for randomised trials: observational study of randomised controlled trials performed by the Radiation Therapy Oncology Group. BMJ 2004;328:22–24. 10.1136/bmj.328.7430.22.

58. Gray R. Why do all systematic reviews have fifteen studies? Nurse Author Ed 2020;30:27–29. 10.1111/nae2.8.

59. Vinkers CH, Lamberink HJ, Tijdink JK, Heus P, Bouter L, Glasziou P, et al. The methodological quality of 176,620 randomized controlled trials published between 1966 and 2018 reveals a positive trend but also an urgent need for improvement. PLoS Biol 2021;19:e3001162. 10.1371/journal.pbio.3001162.

60. Siedler MR, Kawtharany H, Azzam M, Ezgü D, Alshorman A, El Mikati IK, et al. Risk of bias assessment tools often addressed items not related to risk of bias and used numerical scores. J Clin Epidemiol 2025;180:111684. 10.1016/j.jclinepi.2025.111684.

61. Stone J, Gurunathan U, Glass K, Munn Z, Tugwell P, Doi SAR. Stratification by quality induced selection bias in a meta-analysis of clinical trials. J Clin Epidemiol 2019;107:51–59. 10.1016/j.jclinepi.2018.11.015.

62. Jüni P, Witschi A, Bloch R, Egger M. The hazards of scoring the quality of clinical trials for meta-analysis. JAMA 1999;282:1054–1060. 10.1001/jama.282.11.1054.

63. Gordon M, Khudr J, Sinopoulou V, Lakunina S, Rane A, Akobeng A. Quality of reporting inflammatory bowel disease randomised controlled trials: a systematic review. BMJ Open Gastroenterol 2024;11:e001337. 10.1136/bmjgast-2023-001337.

64. Rikos D, Vikelis M, Dermitzakis EV, Soldatos P, Rallis D, Rudolf J, et al. Reporting quality and risk of bias analysis of published RCTs assessing anti-CGRP monoclonal antibodies in migraine prophylaxis: a systematic review. J Clin Med 2024;13:1964. 10.3390/jcm13071964.

65. Viswanathan M, Patnode CD, Berkman ND, Bass EB, Chang S, Hartling L, et al. Recommendations for assessing the risk of bias in systematic reviews of health-care interventions. J Clin Epidemiol 2018;97:26–34. 10.1016/j.jclinepi.2017.12.004.

66. Stadelmaier J, Roux I, Petropoulou M, Schwingshackl L. Empirical evidence of study design biases in nutrition randomised controlled trials: a meta-epidemiological study. BMC Med 2022;20:330. 10.1186/s12916-022-02540-9.

67. Moustgaard H, Jones HE, Savović J, Clayton GL, Sterne JA, Higgins JP, et al. Ten questions to consider when interpreting results of a meta-epidemiological study-the MetaBLIND study as a case. Res Synth Methods 2020;11:260-274. 10.1002/jrsm.1392.

